# Uncovering Multi-Omics Profiles of Population Heterogeneity: A Cluster-Based Bayesian Approach

**DOI:** 10.64898/2025.12.19.25342631

**Authors:** Runye Shi, Ziyang Xiong, Tobias Banaschewski, Gareth J. Barker, Arun L.W. Bokde, Herta Flor, Hugh Garavan, Penny Gowland, Antoine Grigis, Andreas Heinz, Jean-Luc Martinot, Marie-Laure Paillère Martinot, Eric Artiges, Frauke Nees, Dimitri Papadopoulos Orfanos, Luise Poustka, Michael N. Smolka, Sarah Hohmann, Nilakshi Vaidya, Henrik Walter, Robert Whelan, Gunter Schumann, Sylvane Desrivières, Xiaolei Lin, Jianfeng Feng, IMAGEN Consortium

## Abstract

Understanding the heterogeneous nature of genetic effects is critical for advancing our knowledge of the genetic architecture of complex traits and developing personalized management strategies. However, existing methods often rely on pre-specified modifying variables to model this heterogeneity, limiting their ability to capture effects driven by complex or unobserved factors. Here, we propose MOCHA (Multi-Omics Clustering for Heterogeneous Association), a novel Bayesian analytical paradigm that identifies latent population subgroups with distinct genetic effects directly from multi-omics data, without requiring a priori variable specification. Extensive simulations confirm that MOCHA accurately identifies the underlying clustering structure, demonstrates superior performance in identifying and ranking features with cluster-specific effects, and provides reliable parameter estimates. Applying MOCHA to genomic and transcriptomic data from the IMAGEN study, we identified two distinct neurodevelopmental clusters associated with adolescent inhibitory control. Post-hoc characterization of these clusters provided novel insights into the mechanisms of brain plasticity, demonstrating the method’s practical utility and interpretability.

## Introduction

The growing emphasis on precision medicine, driven by the complex interplay between genes and environments, requires a deeper understanding of heterogeneous genetic effects among different subpopulations^1^. This variation arises from multiple regulatory checkpoints across different omics levels in the gene-to-trait pathway^2^. These range from the dynamic nature and plasticity of epigenetic regulation influencing gene expression levels^3^, to post-transcriptional and post-translational processes such as micro-RNA regulation^4^ and context-dependent activation of proteins^5^ through modifications and interactions. Consequently, individuals’ distinct environmental exposure processes, along with their genetic and pathophysiological states, could all contribute to the variations in individual genetic effects^6^. For example, adverse childhood experiences could amplify the genetic susceptibility to bipolar disorder^7^. Similarly, clinically normal individuals carrying APOE ε4 allele have been shown to experience faster short-term cognitive decline in the presence of high β-amyloid accumulation^8^. Dissecting these heterogeneous effects not only offers mechanistic insights into the genetic architecture of complex traits^9^, but also informs disease prevention and management strategies^10^.

Existing research primarily focused on quantifying the prespecified heterogeneity of genetic effects on a multiplicative scale. For example, genome-wide interaction scan augmented the additive genetic model by including an interaction term between a specific genetic variant and a potential environmental modifier^11,12^. Building upon this, methods like BV-LDER-GE^13^ and the MR-G×E^14^ were developed to improve statistical power by incorporating summary statistics of marginal genetic effects. Other methods focused on assessing how overall genetic architecture can be modulated, for instance, through polygenic score-by-environment interactions^15^ or by partitioning trait variation based on genetic interactions^16^. While these studies were confined to a single modifying variable, recent studies such as StructLMM^17^ and GENIE^18^ extended the analytic framework to multidimensional scenarios by modeling environmental similarities shared among individuals or assuming the covariance structure between genetic and environmental factors. Meanwhile, LEMMA^19^ adopted a dimensionality reduction strategy, deriving a single linear combination of observed environments to capture the primary axis of interaction. However, all these methods relied on pre-specified modifying variables, limiting their ability to identify varying genetic effects underlying population heterogeneities.

Here, we develop a multi-omics clustering method for heterogeneous association (MOCHA), a novel data-driven analytical paradigm that does not require a priori specified modifying variables. Specifically, heterogeneous effects of multi-omics features were modeled using a latent cluster structure, which was subsequently linked to the heterogeneity of phenotypic variations among the population. Extensive simulation studies were carried out to validate the performance of MOCHA under different scenarios. Finally, MOCHA was applied to the adolescent inhibitory control in the IMAGEN dataset with genomic and transcriptomic data, identifying two neurodevelopmental clusters.

The use of multi-omics data provided a more comprehensive perspective on the mechanisms of modulated genetic effects in brain plasticity, thereby demonstrating the practical interpretability of our method. Meanwhile, the post-hoc characterization of the identified clusters helped to contextualize these modified effects, suggesting the presence of potential underlying interactions.

## Results

### Overview of the MOCHA model

A graphical overview of MOCHA was presented in Fig. 1. MOCHA is an integrative analytical framework that aims to bridge multi-omics features and heritable phenotypes by specifying latent population clusters with distinct associations between omics features and phenotypes. It allows us to identify multi-omics features ***X*** that contribute to population clustering by adopting a flexible mixture of Gaussian distributions for the heritable phenotype *Y*. Assuming that there are *K* latent clusters, the model is formulated as

**Fig. 1.**
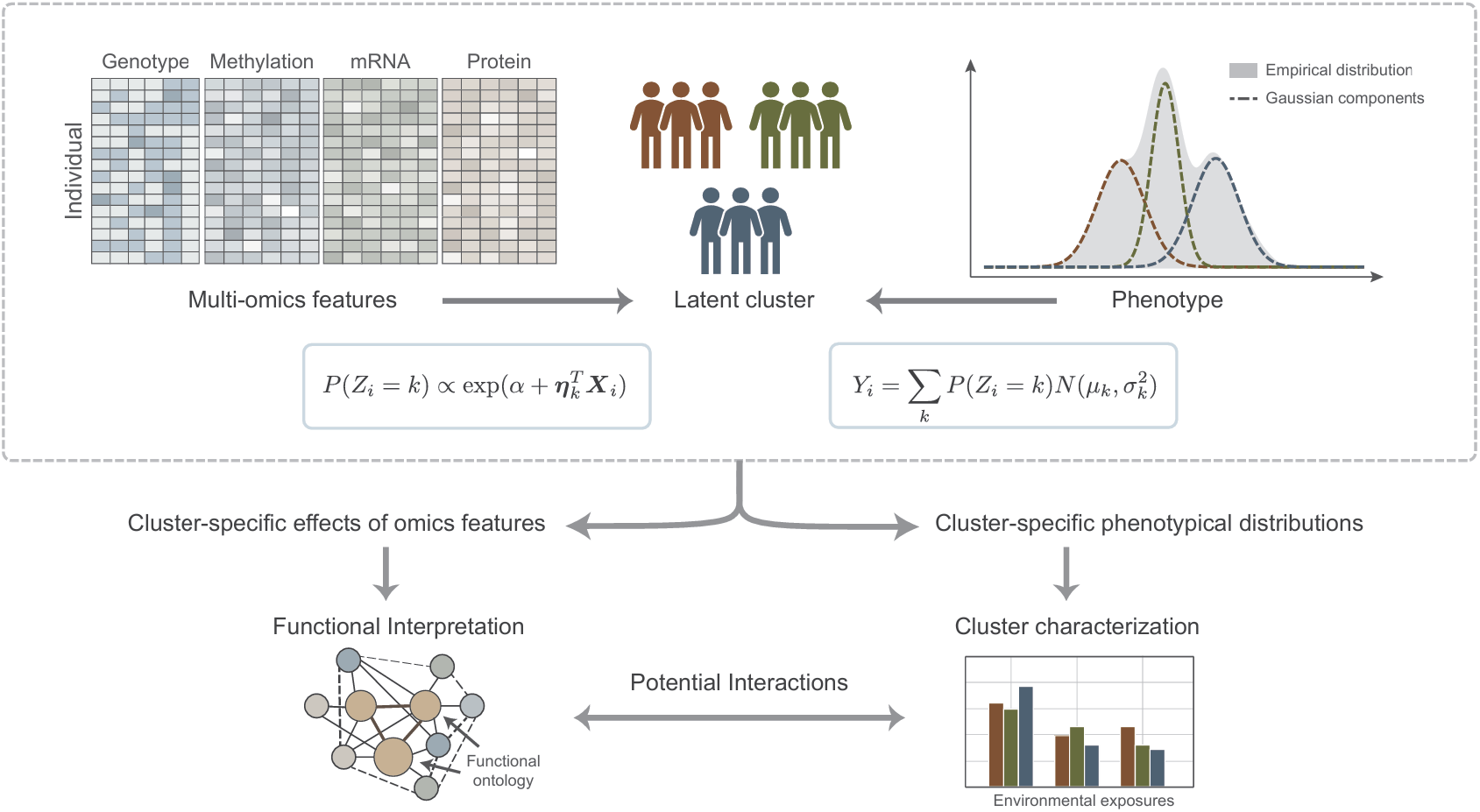
Graphical overview of MOCHA. The method partitions participants into latent clusters by modeling a heritable phenotype as a Gaussian mixture, where cluster assignment is driven by heterogeneous effects of multi-omics features. This enables downstream functional annotation to identify modified biological processes and cluster characterization to define potential modifiers.

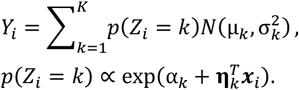

Here, *Z*_*i*_ represents the cluster assignment for participant *i, μ*_*k*_ and 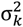 denote the cluster-specific distributional parameters, while *α*_*k*_ and ***η***_*k*_ are the cluster-specific intercept and feature coefficients, respectively. These latent clusters simultaneously reflect both heterogeneity in feature-to-phenotype associations and distinct patterns in phenotypic distributions.

We leveraged a Bayesian Markov Chain Monte Carlo (MCMC) method for parameter estimation. To ensure the identifiability of the mixture components, a preliminary *k*-means clustering was employed to inform the priors of component distributional parameters. To address the major challenge posed by high-dimensional omics features in model fitting and estimation, we assumed that only a few omics features are involved in cluster-specific effects. This was achieved by utilizing a group sparsity structure at the omics level, implying that an omics feature is either selected to have varied influences or not selected with a uniform effect. Specifically, a modified hierarchical prior was applied to feature coefficients, incorporating both global and feature-specific local shrinkage,

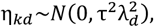

where *k* and *d* denote the cluster and feature indices respectively. While the global shrinkage parameter *τ* could serve as an indicator of the expected proportion of selected features^20^, the magnitude of the local shrinkage parameter *λ*_*d*_ provides a measure of the heterogeneity in feature coefficients across different clusters.

Meanwhile, recognizing that the absolute magnitude of *λ* in the MCMC method could be context-dependent and thus not directly comparable between studies, we further developed a qualitative standard by modeling the threshold for important feature selection^21^ as a linear function of the global shrinkage parameter *τ* (Supplementary Note). This assumption allows the selection threshold to adapt to the overall sparsity of the model. For implementation, we partitioned all features into three credible sets: the large one indicating important features with likely heterogeneous effects, the medium one representing potentially ambiguous ones, and the small one containing unimportant features with likely homogeneous effects.

In addition, to improve computational efficiency, we also implemented an Expectation-Maximization (EM) algorithm for preliminary feature screening. Within the EM model, the selection of cluster-specific omics features was performed using group LASSO, which accommodates different penalty choices for various feature modalities.

A comprehensive mathematical description of MOCHA was presented in the Methods section, with computational implementation details provided in the Supplementary Note.

### MOCHA accurately identifies latent population clusters and cluster-specific multi-omics features in simulation studies

Multi-omics (genotype and transcriptomic profiles) and phenotypic data were simulated to evaluate the performance of MOCHA under multiple scenarios: 1) total sample sizes were set to be 200 or 500, representing high/low feature-to-sample ratios; 2) number of latent population clusters was set to be 3 or 5; 3) modality size ratio was set to be 1:1, 5:1 or 10:1; 4) distribution ratio of true signals across modalities was set to be 1:1 or 5:1. The number of features was set at 1,000. Additive coding genotype data with a minor allele frequency > 0.05 were generated using Plink 1.90, and mRNA expressions were sampled from independent standard normal distributions. The proportion of cluster-specific omics features was fixed at 0.02. For cluster-specific omics features, their effects on each cluster were randomly drawn from *K*non-overlapping uniform distributions, each with a length of 2 and separated by a constant gap of 1; for cluster-constant omics features, effects were sampled from a standard uniform distribution.

To determine the optimal number of latent population clusters, the leave-one-out cross-validated log-likelihood (LOO-CV) for the MCMC approach and the log-likelihood calculated from 5-fold cross-validation for the EM estimation were used as the evaluation metrics. Across all simulation configurations, both the MCMC and EM methods were able to correctly identify the true number of clusters (Fig. 2A and Supplementary Figs. 1-2). Taking a specific scenario as an example (K=5, N=200, modality size ratio and true signal distribution ration being 1:1), omics features with true cluster-specific effects yielded larger estimated *λ*_*MAP*_ values compared to cluster-constant features (Fig. 2B), and *λ*_*MAP*_ also showed excellent discriminatory power, achieving a high Area Under the ROC Curve (AUC) of 0.98 (Fig. 2C). This finding confirmed the ability of the local shrinkage parameter in ranking the feature importance and its utility in distinguishing true signals.

**Fig. 2.**
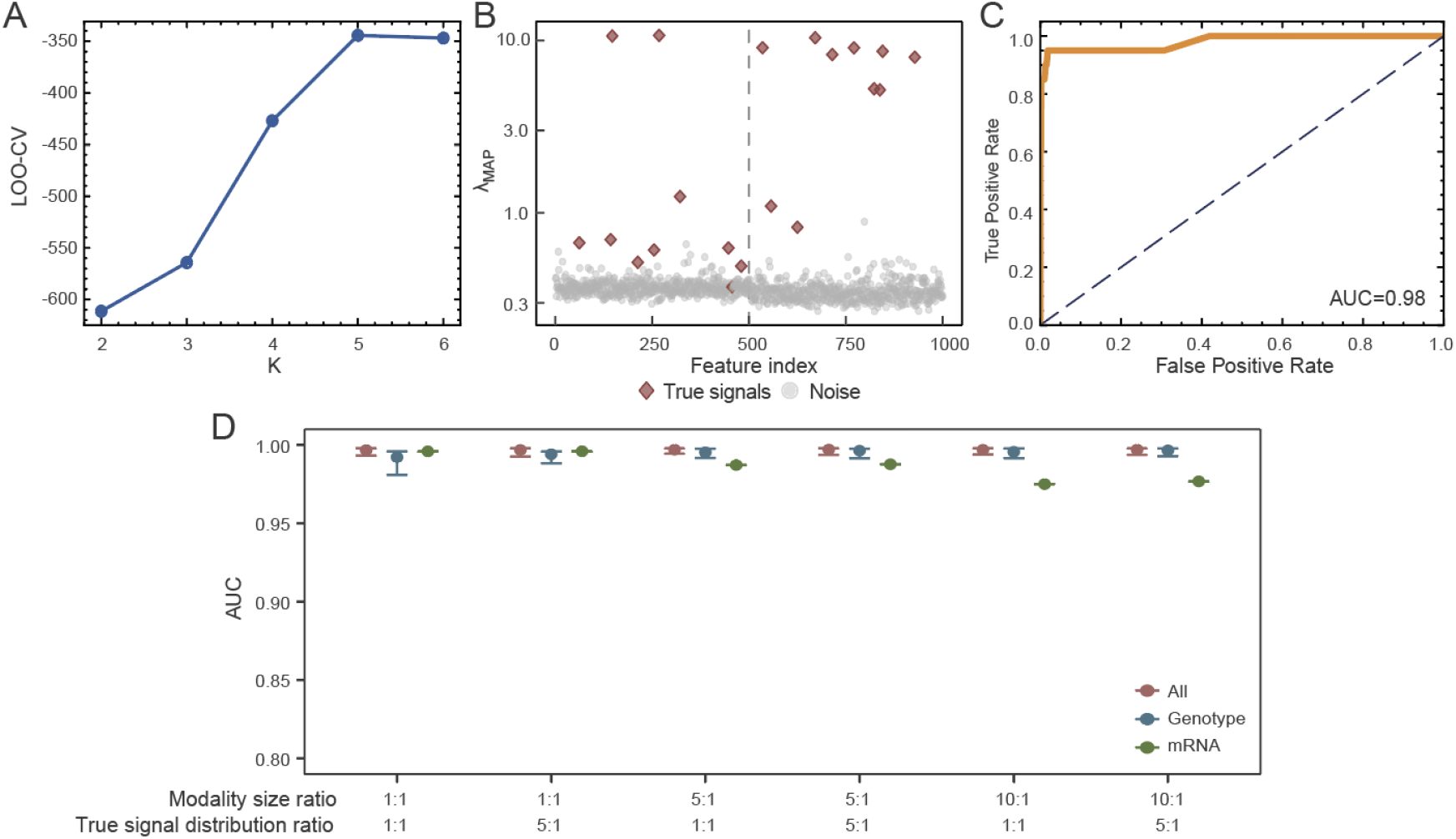
The performance of MCMC implementation on simulated datasets. A-C. Illustration of MCMC algorithm performance on a representative simulated dataset, where K=5, N=200 and modality size ratio/true signal distribution ratio=1:1. A. MCMC approach accurately identified the true number of clusters based on the leave-one-out cross-validation (LOO-CV) estimates. B. True important features and null features were clearly separated by the posterior estimates of local shrinkage parameter, which could be used to rank the feature importance. C. High Area Under the ROC Curve (AUC) demonstrated its accurate identification of features with heterogeneous effects. D. Distribution of AUC results over 400 trials under varying modality size and signal distribution ratios (genotype:mRNA). Each dot represents the mean AUC, with the 5th and 95th percentiles indicated by the lower and upper horizontal lines. Although AUC is proportional to the modality size, the overall performance in feature selection remains generally robust.

To assess the robustness of our algorithm’s performance in feature selection, we independently replicated each configuration 100 times, with the total number of features set to 500. When the number of features was equal for both modalities, the average AUC for mRNA was slightly higher than that for genotype. In contrast, as the proportion of genotype features increased, the AUC for genotype became significantly higher, and this performance difference between the two modalities increased with the increasing ratio (Fig. 2D). Notably, these AUC differences were not influenced by the distribution ratio of true signals across modalities. Despite these performance changes, it is crucial to emphasize that the overall performance remained highly robust, with the AUC consistently exceeding 0.92 across all configurations (Supplementary Table 1), demonstrating a strong and reliable signal identification capability. Meanwhile, for the cluster-specific effects, the 95% HDIs achieved an average coverage rate of 99.36%, suggesting that our algorithm could effectively reflect the magnitudes of the true cluster-specific coefficients (Supplementary Fig. 3).

Next, to compare the cluster-specific omics feature selection between MCMC and EM methods, we established a fixed threshold of *λ*_*MAP*_>0.52 to serve as the selection criterion for cluster-specific features. This threshold was defined as the median of the optimal Youden’s indices derived from AUC calculations (Supplementary Fig. 4), enabling the computation of sensitivity and specificity from the MCMC posterior estimates. Although both algorithms showed improved identification performance with larger number of clusters and sample sizes, the MCMC approach demonstrated greater robustness (Fig. 3A-B and Supplementary Table 2). Compared to EM, MCMC demonstrated well-balanced sensitivity and specificity in feature selection, and at the same time maintained well-controlled false discovery rates across all simulation settings.

**Fig. 3.**
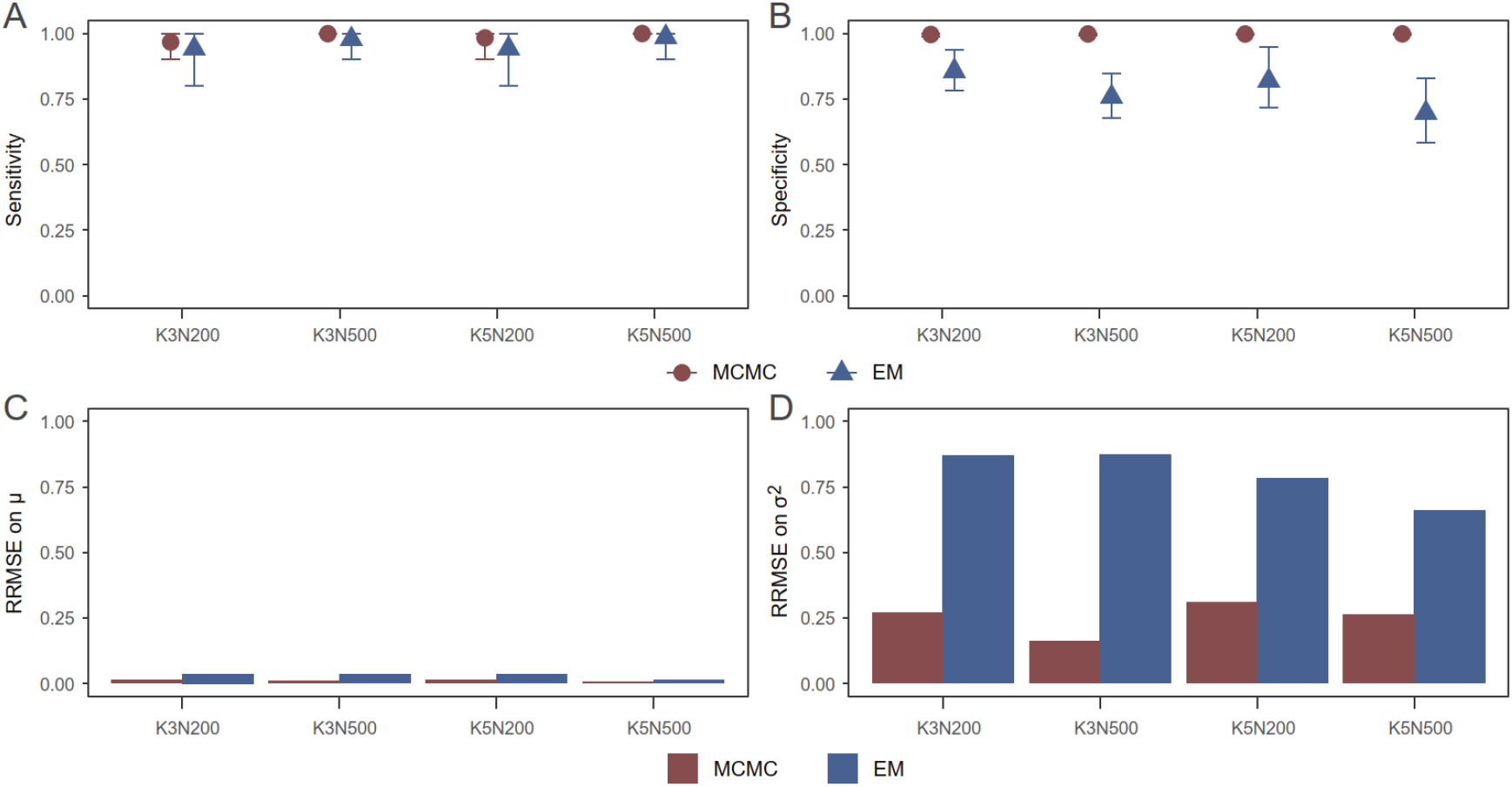
Comparison of MCMC and EM results on simulation datasets. A threshold of 0.52 was used as the inclusion criterion for variable selection in the MCMC algorithm. A-B. Performance in identifying true signals across 600 trials under varying true number of clusters and sample sizes. Each dot represents the mean AUC, with the 5th and 95th percentiles indicated by the lower and upper horizontal lines. Both algorithms show improved identification performance as number of clusters (K) and sample size (N) increase. However, compared to EM, MCMC consistently achieving superior signal identification and well-controlled false discovery rates. C-D. Relative Root Mean Squared Error (RRMSE) of different cluster distribution parameters from 600 trials under varying true number of clusters and sample sizes. RRMSE was calculated for each parameter under different configurations and then summarized by taking the mean. Compared to EM, MCMC shows better parameter estimation performance, especially for variation parameters.

We further validated this framework across a range of simulations, varying the proportion of true important features from 0.01 to 0.05 with increments of 0.005 and setting the population size to either 200 or 500. This approach effectively separated important features (Supplementary Fig. 5 and Supplementary Table 3). The large credible sets defined by features with *λ*_*MAP*_ above the predicted upper thresholds were highly enriched with true important features (proportion 0.81±0.10), while the small set defined by features with *λ*_*MAP*_ below the predicted lower thresholds filtered them out (proportion 0.20±0.07).

Finally, we evaluated the performance of MOCHA in estimating the cluster-specific distributional parameters for candidate phenotypes. Both MCMC and EM methods performed well in estimating the cluster-specific means (Fig. 3C). In comparison, MCMC showed remarkably superior performance in estimating the variances (Fig. 3D and Supplementary Table 4), although we noted its estimates tended to increase as within-cluster sample size decreased, which was not observed with the EM algorithm. This finding highlighted the advantage of MCMC in providing a more accurate characterization of the underlying cluster structure.

### MOCHA identifies two latent adolescent clusters underlying different omics-to-SSRT associations

We applied MOCHA to examine the heterogeneous associations between multi-omics features and inhibitory control measured by stop signal reaction time (SSRT) among adolescents from the IMAGEN cohort (Fig. 4A). After clumping for genotype (MAF>0.05) and correcting batch effects in mRNA expression, a total of 487 adolescents with 96,767 clumped genotypes and 34,833 mRNAs were included in the analysis.

**Fig. 4.**
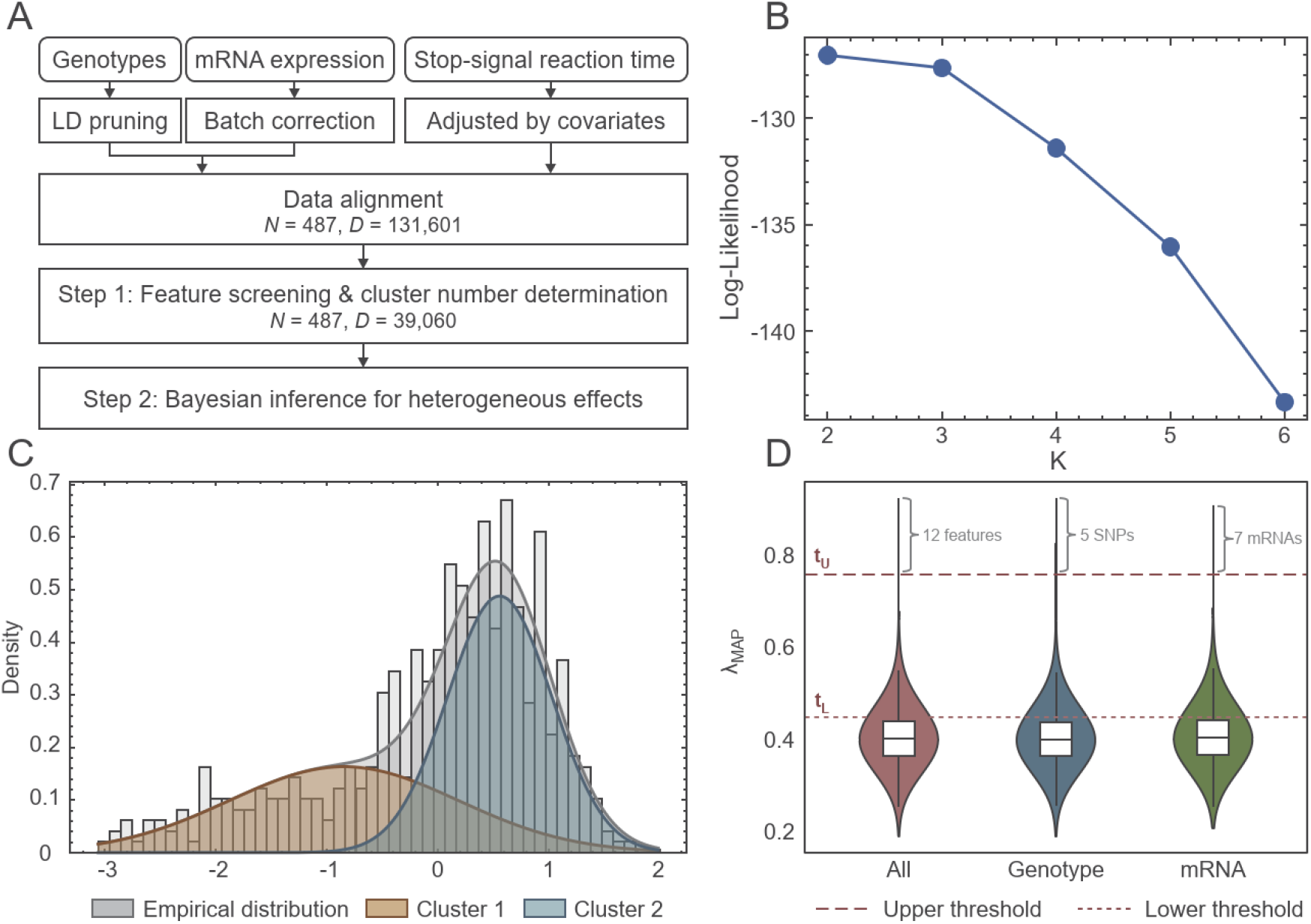
Application of MOCHA in IMAGEN study. A. The workflow of data processing and analysis pipeline using IMAGEN data. LD, linkage disequilibrium. B. Two clusters were identified by the EM method based on the maximum 5-fold cross-validated log-likelihood. C. The empirical phenotype distribution is well-approximated by a two-component Gaussian mixture model. D. The distribution of the posterior estimates for local shrinkage parameters. A threshold-based approach identifies 12 key features (5 SNPs and 7 mRNAs) in the large credible set.

A two-step analytical approach was leveraged to improve computational efficiency. During the first step, we used EM to determine the optimal number of latent clusters and pre-screen both the cluster-specific and cluster-constant omics features. As shown in Fig. 4B, two adolescent clusters were identified using cross-validated log-likelihood, where cluster 1 showed low and more dispersed SSRT (indicating better inhibitory control) and cluster 2 showed high and less variable SSRT (indicating poor inhibitory control). Fig. 4C showed that the bimodal Gaussian mixture provided a good fit for the observed SSRT in the overall adolescent population. The initial prescreening retained a total of 39,060 features (29.68%), comprising 23,249 SNPs (24.03% of all SNPs) and 15,812 mRNAs (45.40% of all mRNAs). During the second step, we used MCMC to refine feature selection and quantify the effect of each selected omics feature

(Supplementary Fig. 6). Based on thresholds trained from the global shrinkage parameter (*τ*_*MAP*_ *= 0.25*), 12 omics features (including 5 SNPs and 7 mRNAs) were identified to have significant cluster-specific effects, while 7,728 features (including 4,385 SNPs and 3,343 mRNAs) fell into the medium credible set, suggesting potentially ambiguous cluster-specific effects (Fig. 4D).

### Omics features with cluster-specific effects pinpoint regulatory networks crucial for adolescent brain plasticity

To elucidate the biological mechanisms of omics features exhibiting distinct roles across clusters, we first mapped both SNPs and mRNAs to the corresponding genes. For SNPs, each variant was assigned to the gene with the highest variant-to-gene score by Open Targets^22^, with each gene weighted by local shrinkage parameter *λ* within each omics modality. Among the 12 omics features identified to have significant cluster-specific effects (Supplementary Data 1), 5 were mapped to genes with known functions for neurodevelopment, including those involved in synapse plasticity (GRID2, MMP2 and EWSR) and neuronal development (MGARP and IRX3).

Gene set enrichment analysis was conducted to further interpret the molecular functions of selected omics features. P-values were calculated using permutation test and Benjamini-Hochberg False Discovery Rate (BH-FDR) was applied to control for multiple testing. From a total of 15,975 gene ontology terms, 529 sets were found to be significant (Supplementary Data 2). A primary functional category related to neuronal structural development and morphogenesis was found based on semantic similarities among the top 15 significant gene sets (Figs. 5A-B), indicating the critical roles of cluster-specific omics features in the maturation of complex neural circuits for higher-order cognitive functions (including inhibitory control) during adolescence^33^. Other functional categories, including the regulation of intracellular signaling pathways (i.e. negative regulation of peptidyl*-*serine phosphorylation), cell migration (i.e. taxis), cellular differentiation (i.e. response to BMP) and fundamental multi-organ development processes (i.e. heart development), stand out as potential molecular functions underlying cluster-specific omics effects.

**Fig. 5.**
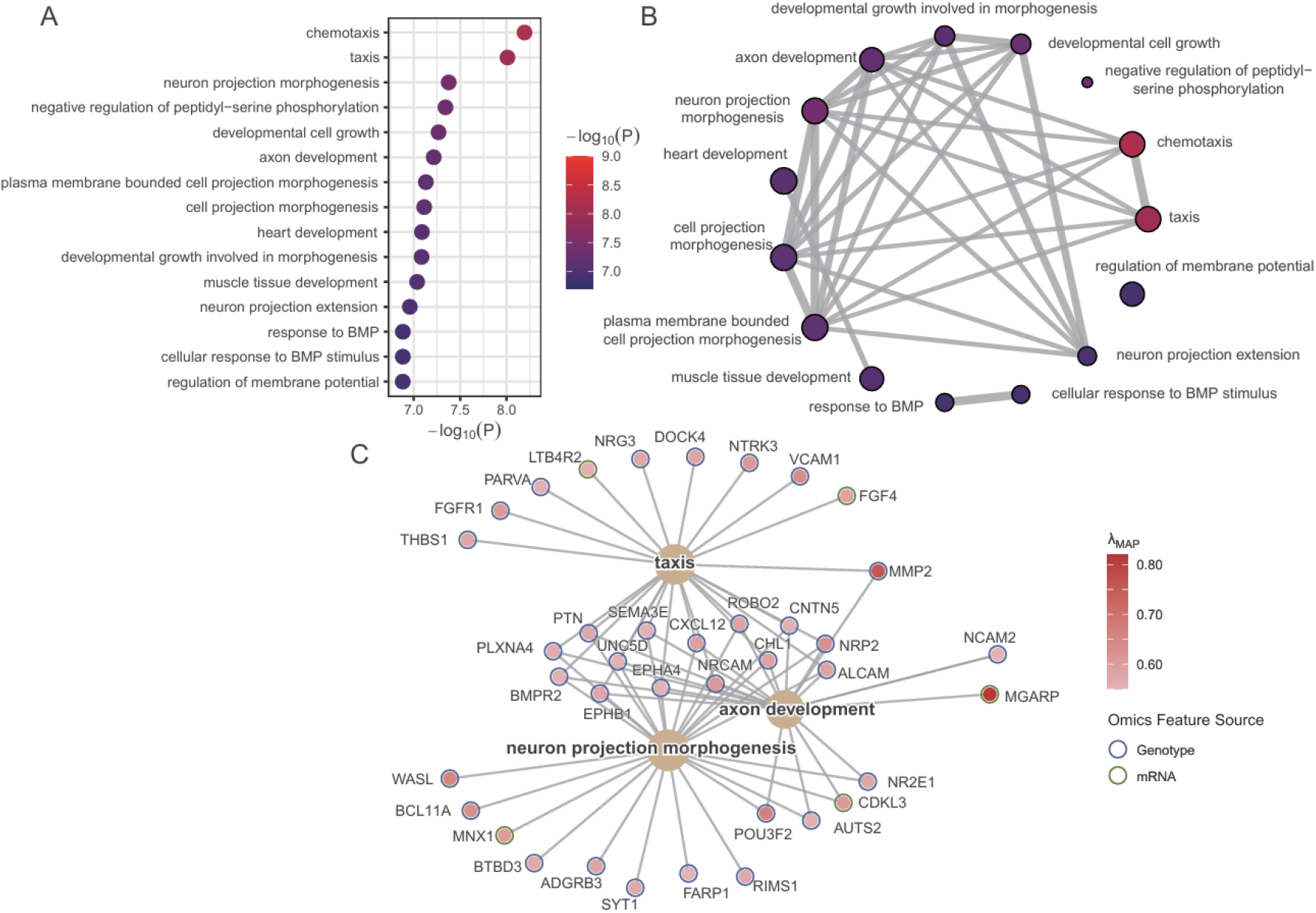
Functional annotation of identified features. A. The top 15 gene sets identified in the gene set enrichment analysis. Genes were weighted by the λ value of the top-ranking feature. P-values were calculated using a permutation test with BH-FDR method for multiple testing correction. B. Semantic similarity network among the top 15 gene sets. Node color indicates the statistical significance as (A). C. Genes with importance weight > 0.55 within the gene sets taxis, neuron projection morphogenesis and axon development.

As an illustrative case, we examined genes with weight larger than 0.55 within the gene sets taxis, neuron projection morphogenesis and axon development (Fig. 5C). High-weighted genes were identified from both omics layers. For example, MMP2 and MGARP were mapped from SNPs and mRNAs respectively. Nevertheless, genomics remained the predominant source for key gene identification. We constructed a gene network using all genes with weights larger than 0.55 (Fig. 6A). Only interactions supported by empirical evidence with a medium confidence score were included. The network highlights a multi-faceted framework governing adolescent brain plasticity, composed of distinct yet interconnected functional modules. It integrates master transcriptional regulators defining developmental programs^29,30,34,35^ (i.e. SOX9, IRX3) with a growth and patterning module guiding cell proliferation and differentiation^36–41^ (i.e. FGFR1, NTRK3, BMPR2). The subsequent wiring of circuits was managed by an axon guidance and cell migration module featuring ephrin–Eph signaling^42^ (i.e. EPHA4, EPIHB1), semaphorin signaling^43,44^ (i.e. SEMA3E, PLXNA4, NRP2) and Slit/Robo signaling^45^ (i.e. ROBO2). The inherent plasticity of these modules underlies the heritability heterogeneity of inhibitory control in adolescent neurodevelopment.

**Fig. 6.**
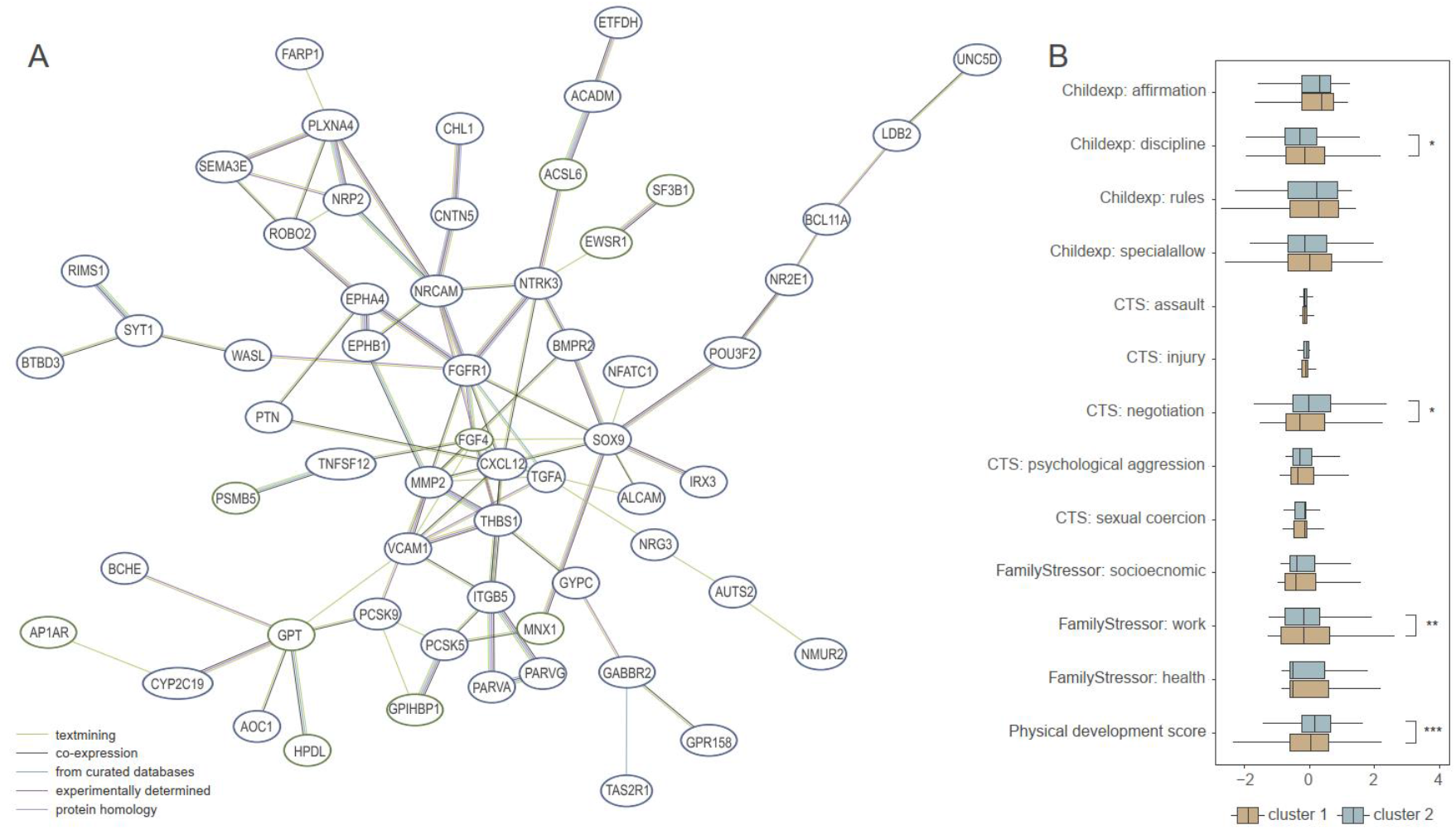
Cluster profiling contextualizes distinct genetic architectures. A. Part of gene interaction network constructed from genes with an importance weight exceeding 0.55. Edges represent known gene-gene interactions, with their color indicating the source of evidence. B. Comparison of physical development and environmental variables between clusters. Significant differences were observed between Cluster 2 and Cluster 1 in measures of parental discipline (d=−0.18, p=0.044), parental conflict negotiation (d=0.19, p=0.027), work-related family stress (d=−0.24, p=0.006), and physical puberty development score (d=0.29, p=0.001). Childexp, child’s experience of family life; FamilyStressor, family stresses; CTS, Conflict Tactics Scale (CTS).

### Cluster profiling contextualizes distinct genetic architectures

We hypothesized that environmental exposures could act as potential modifiers for the cluster-specific omics-SSRT associations. Therefore, we analyzed the differences of puberty development, childhood experiences, parental conflict tactics and family stressors between clusters, adjusting for age, sex and study site. A cross-validated lasso regression was used to select key modifier variables, which were then included in a multivariate logistic regression for statistical inference. Interestingly, cluster 1, with low SSRT and better inhibitory control, showed significantly lower levels of parental discipline (d=−0.18, p=0.044) and work-related family stress (d=−0.24, p=0.006), and higher levels of parental conflict negotiation (d=0.19, p=0.027) and physical puberty development score (d=0.29, p=0.001) (Fig. 6B). This post-hoc characterization of environmental profiles between multi-omics informed adolescent clusters suggested potential environmental modifiers in contextualizing the biological differences shaping heterogeneous inhibitory control.

## Discussion

In this study, we propose MOCHA, a data-driven analytical framework to identify heterogeneous associations between multi-omics features and phenotypes by introducing latent population clusters. A two-stage analytical approach was proposed to enhance computational efficiency, with EM in stage 1 for feature pre-screening and MCMC in stage 2 for feature refinement. Further, a general thresholding strategy using global and local shrinkage parameters was adopted to select cluster-specific omics features. Simulation studies were conducted under various scenarios and demonstrated that MOCHA was able to accurately 1) identify omics features with cluster-specific effects, 2) detect the true numbers of population clusters, and 3) estimate cluster-specific and overall phenotypic distributions, achieving high statistical power while maintaining robust control of false discovery rates. We highlight two key contributions of this new paradigm. Firstly, it does not require a pre-specified set of modifiers in quantifying potential gene-by-environment interactions; rather, it overcomes this limitation by taking on a data-driven approach to capture population heterogeneity underlying differential genomic-phenotypic associations. Secondly, instead of focusing only on SNP-phenotype associations, MOCHA leverages multi-omics features that provide a more comprehensive characterization of potential regulatory mechanisms crucial for understanding the biological functions of each omics modality.

The application of MOCHA to the IMAGEN adolescent cohort yielded novel insights into regulatory pathways and pinpointed potential environmental modifiers contributing to heterogeneous omics-phenotype associations. Our analysis specifically revealed two distinct clusters associated with inhibitory control, as assessed by the SSRT.

These identified cluster-specific features were primarily concentrated in key neurodevelopmental pathways, suggesting a hierarchical regulation of circuit wiring. This is exemplified by several top genes mapped from features in the large credible set. For instance, GRID2 encodes the GluD2 glutamate receptor, a protein highly expressed in the Purkinje cell spines and is essential for synapse organization and central nervous system development^23,24^; MMP2 facilitates the structural plasticity of the nervous system by contributing to axonal outgrowth and regeneration after neuroinflammation^25,26^; MGARP was involved in axonal transport and nervous system development by regulating mitochondrial distribution^27^; IRX3 plays a broad regulatory role in energy homeostasis^28^, neural tube developmental patterning^29^ and neuronal differentiation^30^; EWSR was reported to influence neuronal morphology, dopaminergic signaling and motor function^31^ through its diverse molecular interactions and regulation of mitochondrial function and cellular energy homeostasis^32^.

Beyond these genetic insights, the subsequent environmental characterization of the clusters offers a potential explanation for the observed heterogeneity in omics-to-SSRT associations. Our results suggested that a more relaxed family environment—characterized by less structured parenting and lower family work pressure—along with greater physical maturity, may modulate the gene effects related to brain plasticity, which was subsequently associated with reduced inhibitory control during adolescence. This finding lent support to an interplay between both the developmental readiness and risk social contexts hypotheses^46,47^. Specifically, it points to a developmental asynchrony, where the developmental lag in the prefrontal cortex^48,49^, responsible for decision making and impulse control, makes it difficult for adolescents to manage the complex challenges arising from their physical maturation. This mismatch between physiological and psychological development is then exacerbated within the risk context of an unstructured family environment, which provides insufficient external guidance for their developing self-regulatory capacities.

There are several limitations of this work. First, the proposed analytical framework assumed a Gaussian mixture distribution for the targeted phenotypes, limiting its applicability to discrete phenotypes. This approach could also be suboptimal for quantitative phenotypes with unimodal distributions, where the assumption of Gaussian mixture is prone to overfitting. Future extensions are required to accommodate different types of phenotypical distributions. Second, this method requires individual-level data and is currently unable to handle missing omics features. Since one or more omics modalities could often be missing for certain samples due to cost issues or instrumental limitations, future work on developing built-in strategies for handling missingness in multi-omics features is called for. In addition, while MOCHA was designed for multi-omics features to enhance biological interpretation, the hierarchical structure among omics modalities is not explicitly utilized in the current version. Future research may focus on incorporating multi-omics regulatory networks using information from quantitative trait loci in building an integrated multi-omics framework. Despite these limitations, MOCHA remains a powerful tool for identifying population heterogeneities underlying omics-phenotype associations.

## Methods

### Ethics statement

The IMAGEN study was approved by local ethnical research committees at each research site: King’s College London, University of Nottingham, Trinity College Dublin, University of Heidelberg, Technische Universitat Dresden, Commissariat a l’Energie Atomique et aux Energies Alternatives, and University Medical Center. Informed consents were obtained from all participants and a parent/guardian of each participant under 18 years.

### MOCHA

As shown in Fig. 1, MOCHA assumes that the distribution of a continuous phenotype *Y* follows a finite mixture of Gaussian distributions with *K* components,

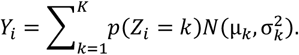

where *Z*_*i*_denotes the cluster assignment for participant *i*. This assignment follows a multinomial distribution, where the probability of participant *i*being assigned to cluster *k* is conditioned on their omics features ***x***_*i*_*∈ ℝ*^*D*^,

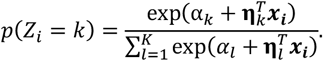

The primary method for fitting the MOCHA model was through Bayesian inference. A plate diagram summarizing the Bayesian model was provided in Supplementary Note. The MCMC sampling was executed via CmdStanPy 1.2.5 with CmdStan 2.36.0, using the No-U-Turn Sampler. We ran 4 parallel chains, generating 1,000 posterior samples for each chain following 1,000 warmup iterations. The target acceptance probability was set to 0.90-0.95. Additionally, we also offered an implementation of the EM algorithm. The penalty parameters were varied across different omics layers. Tree-structured Parzen Estimator sampler was used to perform optimization. Details were provided in the Supplementary Note.

### Simulations

Simulation datasets were constructed to test whether MOCHA could identify clusters with different omics profiles. We assumed that the phenotypic distribution for each of the *K* clusters follows a Gaussian distribution, where the mean was drawn from *N*(*0*,1*0*)and the variance was drawn from *IG*(1,1). To ensure the clusters were identifiable, we constrained the distance between any two adjacent means to be at least four times the larger of their respective standard deviations. Each dataset contains two types of data modalities, additive-coding genotypes and mRNA expression levels. Unlinked genotypes were simulated using Plink 1.90, with minor allele frequencies sampled from *U*(0.05,1.00). The expression data were sampled from independent standard normal distributions. The proportion of important features was set as 0.02. For selected features, their coefficients across different clusters were drawn without replacement from a set of uniform distributions with a length of 2 and a gap of 1. For example, with *K=* 3, the coefficients for a single feature would be drawn from *U*(−4, −2), *U*(−1,1)or *U*(2,4)respectively. Conversely, for null features, the coefficients were set to a single common value across all clusters, which was sampled from a standard uniform distribution. Finally, the cluster assignment for each participant was deterministically set to the cluster with the highest probability, as calculated from their simulated omics features and the generated coefficients.

To evaluate the performance of MOCHA across different configurations, we varied key simulation parameters. The population size was set to 200 or 500 to reflect different sample-to-feature ratios. The number of clusters was set to 3 or 5 to explore varying degrees of population heterogeneity. The genotype-to-mRNA modality size ratio was set to 1, 5 or 10, alongside true signal distribution ratios of 1 or 5, covering a range of signal-to-noise conditions. We designed two simulation scenarios to evaluate MOCHA. In the first scenario, with 1,000 total features, we aimed to determine if MOCHA could accurately identify the true number of clusters under different configurations. In the second scenario, we repeated every configuration 100 times to specifically assess the algorithm’s feature selection and parameter recovery capabilities with a total of 500 features. Regarding the simulations designed to explore thresholding strategies for credible sets, we used the same dataset construction process with the number of clusters constrained to 3 and the proportion of important features ranged from 0.01 to 0.05 with an increment of 0.005.

### Evaluation Metrics

The performance of MOCHA was evaluated in two aspects, the accuracy of cluster parameter recovery and the identification of features with cluster-specific effects. To determine the number of clusters, we employed distinct strategies for each implementation. For the MCMC approach, we compared models using both the LOO-CV, which approximates out-of-sample predictive accuracy. For the EM algorithm, the number of clusters was guided by maximizing the 5-fold cross-validation log-likelihood. In all cases, the elbow method was used to select the optimal model. The accuracy of parameter estimation was quantified using the relative root mean squared error (RRMSE), which provides a scale-invariant measure of the squared error of an estimate relative to the magnitude of the true parameter. For variable selection, since the Bayesian approach does not force coefficients to be exactly zero, we used the MAP estimates of local shrinkage parameters to rank features by importance. The performance of this ranking was then evaluated using the AUC, which was generated by plotting the true positive rate against the false positive rate at 1000 evenly spaced thresholds. In contrast, while the group LASSO penalty performs hard variable selection in the EM algorithm, its variable selection ability was measured by sensitivity and specificity.

### Omics data in the IMAGEN study

We analyzed genotype and gene expression data obtained from the IMAGEN study^50^, a multicenter genetic-neuroimaging project focused on healthy adolescents of European ancestry. Genotypes were assessed in IMAGEN using two genotyping arrays, the Illumina Human610-Quad BeadChip and the Illumina Human660-Quad BeadChip arrays, where QC procedures were applied separately. Non-autosomal variants were removed and SNPs with call rate<95%, minor allele frequency<5%, deviation from Hardy–Weinberg equilibrium (P≤1E-3), or non-autosomal were excluded. Individuals were also excluded due to high genotype failure rate (>5%), divergent ancestry identified by population structure analysis, close relatedness detected by identity-by-state clustering or outlier status determined by principal component analysis (defined as >4 standard deviations from the mean on any of the first 20 PCs). After QC, genotypes from two platforms were merged, with platform-specific SNPs excluded. Then we conducted an imputation using the TOPMed imputation server based on the HapMap3 reference panel^51^, resulting in 5,966,316 SNPs and 1,982 samples. Baseline gene expression data were quantified from whole blood samples using Illumina HumanHT-12 v4 Expression BeadChips, which was then normalized using the mloess method^52^ and log-transformed. ComBat was used to adjust for batch effects in expression data. A total of 34,833 mRNAs were available for 631 participants.

### Data preparation for MOCHA

We used the baseline stop signal reaction time as our phenotype of interest, which was adjusted by age, gender, handedness and site before analysis. Stop signal task is a cognitive test designed to measure inhibitory control^53^, where participants are required to respond to a go signal but stop if a subsequent stop signal appears. SSRT is calculated based on the participant’s reaction time on go trials and their success rate at stopping, with a lower value indicative of better inhibitory control. A total of 487 participants with both omics and cognitive measurements were included.

To handle missing values possibly due to measurement or quality control procedures, we removed SNPs with any missing calls and imputed mRNAs using the corresponding mean expression levels. We also re-filtered the genetic data, retaining only SNPs with a minor allele frequency>0.05. To account for linkage disequilibrium, we further performed pruning on SNPs using PLINK1.9 based on a window size of 5, a step size of 10, and an r^2^ threshold of 0.2. Finally, a total of 96,767 SNPs and 34,833 mRNAs were included in the analysis.

### Cluster identification and profiling

Given that the efficiency of MCMC sampling is sensitive to the number of parameters, we leveraged a two-step approach to improve computational efficiency. We first used the EM algorithm to determine the optimal number of clusters and to perform an initial feature pre-selection. Subsequently, the MCMC algorithm was employed to refine the variable selection and quantify the importance of each feature by the MAP estimates of the local shrinkage parameter *λ*.

To elucidate the modified functional processes between clusters, we mapped both SNPs and mRNAs to genes. Each SNP was assigned to the gene with the highest variant-to-gene score provided by Open Targets^22^ (*genesForVariant()* in otargen-1.1.5 package), while each mRNA was linked to its corresponding gene using the annotation file from array. The importance score of each gene was decided by the *λ*_*MAP*_ value of the top-ranking feature within each omics modality. Genes were standardized by the Entrez ID and were further mapped to the gene ontology. The enrichment analysis was conducted with genes weighted by the importance scores (*gseGO()* in clusterProfiler-4.6.2 package). P-values were obtained via permutation testing and BH-FDR method was used for multiple testing. Semantic similarities between gene sets were calculated using *pairwise_termsim()* in enrichplot-1.18.4 package. Gene-gene interaction network was derived from STRING v12.0^54^. Only interactions with a medium confidence score (score>0.4) and supported by empirical evidence from curated databases, experiments, text mining, co-expression or protein homology sources were included.

To contextualize the regulation process, we examined the physical development and environmental differences between clusters. Physical Development Scale, child’s experience of family life (Childexp) and family stresses (FamilyStressor) from the Development and Well-Being Assessment scale, and Conflict Tactics Scale (CTS) were used. All the measurements were adjusted for age, gender and site. Participants were assigned to clusters based on the maximum probability. Recognizing that this resulted in an imbalanced class distribution, we employed the Synthetic Minority Over-sampling Technique (SMOTE) to balance the data prior to model training (*SMOTE()* in smotefamily-1.4.0 package). Next, a cross-validated LASSO regression was used to select the most informative features (*cv.glmnet()* in glmnet-4.1-9 package). These selected features were then included in a multivariate logistic regression model to assess the statistical significance.

## Data Availability

IMAGEN data can be accessed by email at https://imagen-project.org/ upon application.

## Acknowledgments

This work received support from the following sources: the General Projects of Shanghai Science and Technology Commission (21ZR1405000 [X.L.]), the National Nature Science Foundation of China (No.82304241 [X.L.]), National Key R&D Program of China (No.2018YFC1312904 [J.F.], No.2019YFA0709502 [J.F.]), Shanghai Municipal Science and Technology Major Project (No.2018SHZDZX01 [J.F.], ZJ Lab [J.F.], and Shanghai Center for Brain Science and Brain-Inspired Technology [J.F.]), the 111 Project (No.B18015 [J.F.]), the European Union-funded FP6 Integrated Project IMAGEN (Reinforcement-related behaviour in normal brain function and psychopathology) (LSHM-CT-2007-037286 [G.S.]), the UK Research and Innovation (UKRI) funded UK government’s Horizon Europe funding guarantee (10041392 and 10038599 [G.S.]), the Horizon 2020 funded ERC Advanced Grant ‘STRATIFY’ ((Brain network based stratification of reinforcement-related disorders) (695313 [G.S.]), Human Brain Project (HBP SGA 2, 785907, and HBP SGA 3, 945539 [G.S.]), the Medical Research Council Grant ‘c-VEDA’ (Consortium on Vulnerability Externalizing Disorders and Addictions) (MR/N000390/1 [G.S.]), the National Institute of Health (NIH) (R01DA049238 [G.S.], A decentralized macro and micro gene-by-environment interaction analysis of substance use behavior and its brain biomarkers), the National Institute for Health Research (NIHR) Biomedical Research Centre at South London and Maudsley NHS Foundation Trust and King’s College London, the Bundesministeriumfür Bildung und Forschung (BMBF grants 01GS08152; 01EV0711 [G.S.]; Forschungsnetz AERIAL 01EE1406A, 01EE1406B; Forschungsnetz IMAC-Mind 01GL1745B [G.S.]), the Deutsche Forschungsgemeinschaft (DFG grants SM 80/7-2, SFB 940, TRR 265, NE 1383/14-1 [G.S.]), the Medical Research Foundation and Medical Research Council (grants MR/R00465X/1 and MR/S020306/1 [S.D.]), the National Institutes of Health (NIH) funded ENIGMA (grants 5U54EB020403-05 and 1R56AG058854-01 [S.D.]), NSFC grant 82150710554 and environMENTAL grant 101057429. Further support was provided by grants from: - the ANR (ANR-12-SAMA-0004, AAPG2019 - GeBra [J.-L.M.]), the Eranet Neuron (AF12-NEUR0008-01 - WM2NA; and ANR-18-NEUR00002-01 - ADORe [J.-L.M.]), the Fondation de France (00081242 [J.-L.M.]), the Fondation pour la Recherche Médicale (DPA20140629802 [J.-L.M.]), the Mission Interministérielle de Lutte-contre-les-Drogues-et-les-Conduites-Addictives (MILDECA [J.-L.M.]), the Assistance-Publique-Hôpitaux-de-Paris and INSERM (interface grant [M.-L.P.M.]), Paris Sud University IDEX 2012 [J.-L.M.], the Fondation de l’Avenir (grant AP-RM-17-013 [M.-L.P.M.]), the Fédération pour la Recherche sur le Cerveau; the National Institutes of Health, Science Foundation Ireland (16/ERCD/3797 [R.W.]) and by NIH Consortium grant U54 EB020403 [S.D.], supported by a cross-NIH alliance that funds Big Data Knowledge Centres of Excellence. The funders had no role in study design, data collection and analysis, decision publish or preparation of the manuscript.

## Contributions

R.S., X.L., and J.F. conceptualized the study; R.S. and X.L. designed the analytic approach; S.D. helped in the study design; R.S. implemented the algorithm, interpreted the results and visualized the results; Z. X helped in computational details; R.S. wrote the manuscript; X.L. and J.F. revised the first draft; T.B., G.J.B., A.L.W.B., S.D., H.F., A.G., H.G., P.G., A.H., J.-L.M., M.-L.P.M., E.A., F.N., D.P.O., L.P., S.H, M.N.S., N.V., H.W., R.W., and G.S. were the principal investigators of IMAGEN Consortium; Imaging, genetic and behavioral data in the IMAGEN project were acquired and provided by the IMAGEN Consortium; All authors critically revised the manuscript. X.L. and J.F. contributed equally to this paper.

## References

1 Roberts, M. C., Holt, K. E., Del Fiol, G., Baccarelli, A. A. & Allen, C. G. Precision public health in the era of genomics and big data. Nat Med 30, 1865–1873, doi:10.1038/s41591-024-03098-0 (2024).

2 Motsinger-Reif, A. A. et al. Gene-environment interactions within a precision environmental health framework. Cell Genom 4, 100591, doi:10.1016/j.xgen.2024.100591 (2024).

3 Aguilera, O., Fernandez, A. F., Munoz, A. & Fraga, M. F. Epigenetics and environment: a complex relationship. J Appl Physiol (1985) 109, 243–251, doi:10.1152/japplphysiol.00068.2010 (2010).

4 Hudder, A. & Novak, R. F. miRNAs: effectors of environmental influences on gene expression and disease. Toxicol Sci 103, 228–240, doi:10.1093/toxsci/kfn033 (2008).

5 Devi, S., Chaturvedi, M., Fatima, S. & Priya, S. Environmental factors modulating protein conformations and their role in protein aggregation diseases. Toxicology 465, 153049, doi:10.1016/j.tox.2021.153049 (2022).

6 Baccarelli, A., Dolinoy, D. C. & Walker, C. L. A precision environmental health approach to prevention of human disease. Nat Commun 14, 2449, doi:10.1038/s41467-023-37626-2 (2023).

7 Park, Y. M., Shekhtman, T. & Kelsoe, J. R. Interaction between adverse childhood experiences and polygenic risk in patients with bipolar disorder. Transl Psychiatry 10, 326, doi:10.1038/s41398-020-01010-1 (2020).

8 Mormino, E. C. et al. Amyloid and APOE epsilon4 interact to influence short-term decline in preclinical Alzheimer disease. Neurology 82, 1760–1767, doi:10.1212/WNL.0000000000000431 (2014).

9 Sevilla-González, M. et al. Heterogeneous effects of genetic variants and traits associated with fasting insulin on cardiometabolic outcomes. Nature Communications 16, 2569, doi:10.1038/s41467-025-57452-y (2025).

10 Woodward, A. A., Urbanowicz, R. J., Naj, A. C. & Moore, J. H. Genetic heterogeneity: Challenges, impacts, and methods through an associative lens. Genet Epidemiol 46, 555–571, doi:10.1002/gepi.22497 (2022).

11 Shi, R. et al. Gene-environment interactions in the influence of maternal education on adolescent neurodevelopment using ABCD study. Sci Adv 10, eadp3751, doi:10.1126/sciadv.adp3751 (2024).

12 Ritz, B. R. et al. Lessons Learned From Past Gene-Environment Interaction Successes. Am J Epidemiol 186, 778–786, doi:10.1093/aje/kwx230 (2017).

13 Dong, Z. et al. Incorporating additive genetic effects and linkage disequilibrium information to discover gene-environment interactions using BV-LDER-GE. Genome Biology 26, 332, doi:10.1186/s13059-025-03815-z (2025).

14 Zhu, X. et al. An approach to identify gene-environment interactions and reveal new biological insight in complex traits. Nat Commun 15, 3385, doi:10.1038/s41467-024-47806-3 (2024).

15 Durvasula, A. & Price, A. L. Distinct explanations underlie gene-environment interactions in the UK Biobank. Am J Hum Genet 112, 644–658, doi:10.1016/j.ajhg.2025.01.014 (2025).

16 Yang, J., Lee, S. H., Goddard, M. E. & Visscher, P. M. GCTA: a tool for genome-wide complex trait analysis. Am J Hum Genet 88, 76–82, doi:10.1016/j.ajhg.2010.11.011 (2011).

17 Moore, R. et al. A linear mixed-model approach to study multivariate gene-environment interactions. Nat Genet 51, 180–186, doi:10.1038/s41588-018-0271-0 (2019).

18 Pazokitoroudi, A. et al. A scalable and robust variance components method reveals insights into the architecture of gene-environment interactions underlying complex traits. Am J Hum Genet 111, 1462–1480, doi:10.1016/j.ajhg.2024.05.015 (2024).

19 Kerin, M. & Marchini, J. Inferring Gene-by-Environment Interactions with a Bayesian Whole-Genome Regression Model. Am J Hum Genet 107, 698–713, doi:10.1016/j.ajhg.2020.08.009 (2020).

20 Pas, S. L. v. d., Kleijn, B. J. K. & Vaart, A. W. v. d. The horseshoe estimator: Posterior concentration around nearly black vectors. Electronic Journal of Statistics 8, 2585–2618, doi:10.1214/14-EJS962 (2014).

21 Stéphanie van der, P., Botond, S. & Aad van der, V. Uncertainty Quantification for the Horseshoe (with Discussion). Bayesian Analysis 12, 1221–1274, doi:10.1214/17-BA1065 (2017).

22 Buniello, A. et al. Open Targets Platform: facilitating therapeutic hypotheses building in drug discovery. Nucleic Acids Res 53, D1467–D1475, doi:10.1093/nar/gkae1128 (2025).

23 Allen, J. P. et al. Clinical features, functional consequences, and rescue pharmacology of missense GRID1 and GRID2 human variants. Hum Mol Genet 33, 355–373, doi:10.1093/hmg/ddad188 (2024).

24 Grigorenko, A. P. et al. Neurodevelopmental Syndrome with Intellectual Disability, Speech Impairment, and Quadrupedia Is Associated with Glutamate Receptor Delta 2 Gene Defect. Cells 11, doi:10.3390/cells11030400 (2022).

25 Webber, C. A., Hocking, J. C., Yong, V. W., Stange, C. L. & McFarlane, S. Metalloproteases and guidance of retinal axons in the developing visual system. J Neurosci 22, 8091–8100, doi:10.1523/JNEUROSCI.22-18-08091.2002 (2002).

26 Yong, V. W., Power, C., Forsyth, P. & Edwards, D. R. Metalloproteinases in biology and pathology of the nervous system. Nat Rev Neurosci 2, 502–511, doi:10.1038/35081571 (2001).

27 Jia, L., Liang, T., Yu, X., Ma, C. & Zhang, S. MGARP regulates mouse neocortical development via mitochondrial positioning. Mol Neurobiol 49, 1293–1308, doi:10.1007/s12035-013-8602-8 (2014).

28 Dou, Z., Son, J. E. & Hui, C. C. Irx3 and Irx5 - Novel Regulatory Factors of Postnatal Hypothalamic Neurogenesis. Front Neurosci 15, 763856, doi:10.3389/fnins.2021.763856 (2021).

29 Briscoe, J., Pierani, A., Jessell, T. M. & Ericson, J. A homeodomain protein code specifies progenitor cell identity and neuronal fate in the ventral neural tube. Cell 101, 435–445, doi:10.1016/s0092-8674(00)80853-3 (2000).

30 Szu, J., Wojcinski, A., Jiang, P. & Kesari, S. Impact of the Olig Family on Neurodevelopmental Disorders. Front Neurosci 15, 659601, doi:10.3389/fnins.2021.659601 (2021).

31 Yoon, Y. et al. Genetic Ablation of EWS RNA Binding Protein 1 (EWSR1) Leads to Neuroanatomical Changes and Motor Dysfunction in Mice. Exp Neurobiol 27, 103–111, doi:10.5607/en.2018.27.2.103 (2018).

32 Lee, J. et al. EWSR1, a multifunctional protein, regulates cellular function and aging via genetic and epigenetic pathways. Biochim Biophys Acta Mol Basis Dis 1865, 1938–1945, doi:10.1016/j.bbadis.2018.10.042 (2019).

33 Constantinidis, C. & Luna, B. Neural Substrates of Inhibitory Control Maturation in Adolescence. Trends Neurosci 42, 604–616, doi:10.1016/j.tins.2019.07.004 (2019).

34 Molofsky, A. V. et al. Expression profiling of Aldh1l1-precursors in the developing spinal cord reveals glial lineage-specific genes and direct Sox9-Nfe2l1 interactions. Glia 61, 1518–1532, doi:10.1002/glia.22538 (2013).

35 Jo, A. et al. The versatile functions of Sox9 in development, stem cells, and human diseases. Genes Dis 1, 149–161, doi:10.1016/j.gendis.2014.09.004 (2014).

36 Ohkubo, Y., Uchida, A. O., Shin, D., Partanen, J. & Vaccarino, F. M. Fibroblast growth factor receptor 1 is required for the proliferation of hippocampal progenitor cells and for hippocampal growth in mouse. J Neurosci 24, 6057–6069, doi:10.1523/JNEUROSCI.1140-04.2004 (2004).

37 Tome, D., Dias, M. S., Correia, J. & Almeida, R. D. Fibroblast growth factor signaling in axons: from development to disease. Cell Commun Signal 21, 290, doi:10.1186/s12964-023-01284-0 (2023).

38 Otnaess, M. K. et al. Evidence for a possible association of neurotrophin receptor (NTRK-3) gene polymorphisms with hippocampal function and schizophrenia. Neurobiol Dis 34, 518–524, doi:10.1016/j.nbd.2009.03.011 (2009).

39 Nikoletopoulou, V. et al. Neurotrophin receptors TrkA and TrkC cause neuronal death whereas TrkB does not. Nature 467, 59–63, doi:10.1038/nature09336 (2010).

40 Bartkowska, K., Paquin, A., Gauthier, A. S., Kaplan, D. R. & Miller, F. D. Trk signaling regulates neural precursor cell proliferation and differentiation during cortical development. Development 134, 4369–4380, doi:10.1242/dev.008227 (2007).

41 Nasim, M. T. et al. BMPR-II deficiency elicits pro-proliferative and anti-apoptotic responses through the activation of TGFbeta-TAK1-MAPK pathways in PAH. Hum Mol Genet 21, 2548–2558, doi:10.1093/hmg/dds073 (2012).

42 Kania, A. & Klein, R. Mechanisms of ephrin-Eph signalling in development, physiology and disease. Nat Rev Mol Cell Biol 17, 240–256, doi:10.1038/nrm.2015.16 (2016).

43 Lu, D., Shang, G., He, X., Bai, X. C. & Zhang, X. Architecture of the Sema3A/PlexinA4/Neuropilin tripartite complex. Nat Commun 12, 3172, doi:10.1038/s41467-021-23541-x (2021).

44 Suto, F. et al. Plexin-a4 mediates axon-repulsive activities of both secreted and transmembrane semaphorins and plays roles in nerve fiber guidance. J Neurosci 25, 3628–3637, doi:10.1523/JNEUROSCI.4480-04.2005 (2005).

45 Gore, B. B. et al. Roundabout receptor 2 maintains inhibitory control of the adult midbrain. Elife 6, doi:10.7554/eLife.23858 (2017).

46 Lewis, M., Miller, S. M. & Sameroff, A. J. Handbook of developmental psychopathology. (1990).

47 Pfeifer, J. H. & Allen, N. B. Puberty Initiates Cascading Relationships Between Neurodevelopmental, Social, and Internalizing Processes Across Adolescence. Biol Psychiatry 89, 99–108, doi:10.1016/j.biopsych.2020.09.002 (2021).

48 Juraska, J. M. The last stage of development: The restructuring and plasticity of the cortex during adolescence especially at puberty. Dev Psychobiol 66, e22468, doi:10.1002/dev.22468 (2024).

49 Ordaz, S. J., Foran, W., Velanova, K. & Luna, B. Longitudinal growth curves of brain function underlying inhibitory control through adolescence. J Neurosci 33, 18109–18124, doi:10.1523/JNEUROSCI.1741-13.2013 (2013).

50 Mascarell Maricic, L. et al. The IMAGEN study: a decade of imaging genetics in adolescents. Mol Psychiatry 25, 2648–2671, doi:10.1038/s41380-020-0822-5 (2020).

51 International HapMap, C. et al. Integrating common and rare genetic variation in diverse human populations. Nature 467, 52–58, doi:10.1038/nature09298 (2010).

52 Sasik, R., Calvo, E. & Corbeil, J. Statistical analysis of high-density oligonucleotide arrays: a multiplicative noise model. Bioinformatics 18, 1633–1640, doi:10.1093/bioinformatics/18.12.1633 (2002).

53 Bari, A. & Robbins, T. W. Inhibition and impulsivity: behavioral and neural basis of response control. Prog Neurobiol 108, 44–79, doi:10.1016/j.pneurobio.2013.06.005 (2013).

54 Szklarczyk, D. et al. The STRING database in 2023: protein-protein association networks and functional enrichment analyses for any sequenced genome of interest. Nucleic Acids Res 51, D638–D646, doi:10.1093/nar/gkac1000 (2023).

